# Modeling a traffic light warning system for acute respiratory infections

**DOI:** 10.1101/2022.12.16.22283591

**Authors:** Saul Diaz-Infante, M. Adrian Acuña-Zegarra, Jorge X. Velasco-Hernández

## Abstract

The high morbidity of acute respiratory infections constitutes a crucial global health burden. In particular, for SARS-CoV-2, non-pharmaceutical intervention geared to enforce social distancing policies, vaccination, and treatments will remain an essential part of public health policies to mitigate and control disease outbreaks. However, the implementation of mitigation measures directed to increase social distancing when the risk of contagion is a complex enterprise because of the impact of NPI on beliefs, political views, economic issues, and, in general, public perception. The way of implementing these mitigation policies studied in this work is the so-called traffic-light monitoring system that attempts to regulate the application of measures that include restrictions on mobility and the size of meetings, among other non-pharmaceutical strategies. Balanced enforcement and relaxation of measures guided through a traffic-light system that considers public risk perception and economic costs may improve the public health benefit of the policies while reducing their cost. We derive a model for the epidemiological traffic-light policies based on the best response for trigger measures driven by the risk perception of people, instant reproduction number, and the prevalence of a hypothetical acute respiratory infection. With numerical experiments, we evaluate and identify the role of appreciation from a hypothetical controller that could opt for protocols aligned with the cost due to the burden of the underlying disease and the economic cost of implementing measures. As the world faces new acute respiratory outbreaks, our results provide a methodology to evaluate and develop traffic light policies resulting from a delicate balance between health benefits and economic implications.

## 1. Introduction

Acute respiratory infections (ARI) are an important health burden for all human populations around the globe. These infections have a higher impact (higher morbidity or mortality) on the very young or the very old individuals, with an estimated 156 million infections every year and an important associated mortality rate [1]. According to these authors, the most common ARIs correspond to the respiratory syncytial virus (RSV), human metapneumovirus, rhinovirus/enterovirus, influenza viruses, adenovirus, coronavirus, *Streptococcus pneumoniae*, and *Mycoplasma pneumoniae*. Associated with the circulation of these pathogens, the seasonal emergence of outbreaks is another characteristic of ARIs. A recent study in China [2] reported a seasonal pattern mainly driven by RSV and influenza viruses with influenza virus, respiratory syncytial virus, and human rhinovirus as the three leading viral pathogens and *Streptococcus pneumoniae, Mycoplasma pneumoniae* and *Klebsiella pneumoniae* the three most important bacterial pathogens. Currently, there is no vaccine or effective antiviral treatment against RSV, but several antivirals and vaccines are available for influenza [1]. In view of these differences in the control of ARIs, it would be expected that non-pharmaceutical interventions (NPIs), geared to enforce social distancing policies, in combination with vaccination and treatments will remain an important part of public health policies that seek to mitigate and control disease outbreaks. As an illustrative example of the dynamics that may occur in ARIs, we show in Figure 1 the time series of the COVID-19 pandemic, registered in Mexico for one year of the pandemic from March 23, 2020) until vaccines started to be introduced. The complex but periodic dynamics that can be appreciated is likely the result of the interaction of the natural dynamics of the disease, the implementation of NPIs and weather variability.

**Figure 1:**
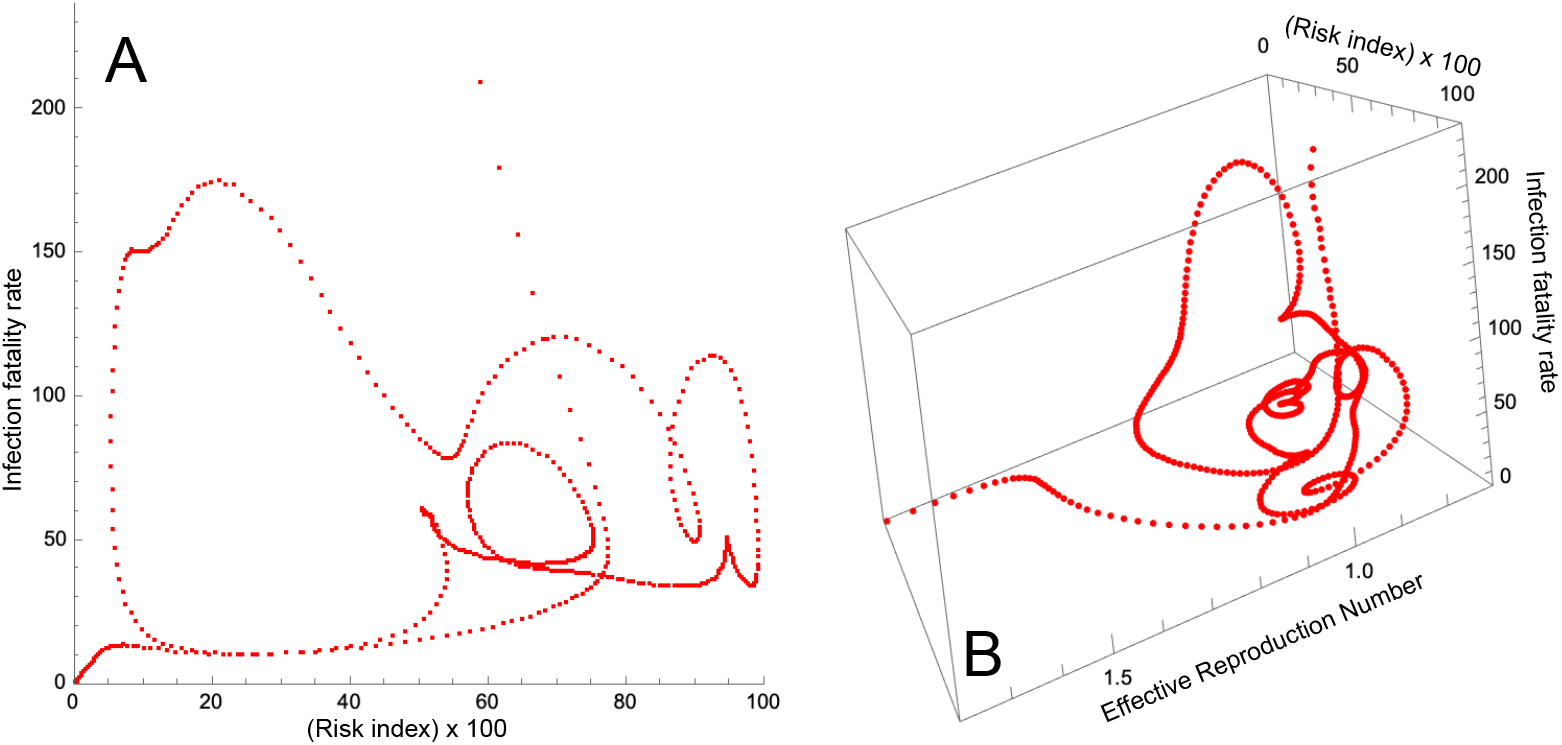
Epidemic orbits for the first 540 days of the pandemic. A) Risk index-IFR plane for the state of Queretaro. The *x*-axis is the reproductive number and the *y*-axis is the IFR. B) *R*_*t*_, *C*_*k*_(*t*), and IFR 3D orbit for Mexico City (*x, y* and *z* axes, respectively).

SARS-CoV-2 has joined the group of ARIs that will be cocirculating in the future around the World; it is also an example of how NPIs were implemented to control the disease. All over the world, governmental institutions were in charge of defining criteria for the lifting or implementation of NPIs based on indicators related to risk perception and economic feasibility. In Mexico and New Zealand, for example, the policies of mitigation and control were integrated into a traffic light system to regulate social distancing that included partial or full closures of schools, or enforced limited capacity for events, public transportation, and so forth, depending upon a series of indicators among which the most important were the state of the epidemic as measured by prevalence, the value of the reproduction number, the number of hospitalizations, and deaths. However, the effectiveness of implementation of mitigation measures directed to increase social distancing, when the risk of contagion is perceived as large, has been controversial. For example, [3], points out that lockdowns have not a clearly identified effect on epidemic control with its efficacy depending very much on the target population, its level of economic development, its confidence in government, and other factors.

Due to the above, the following question arises: is it possible to design an indicator based on the population dynamics of the disease, that coupled to NPI interventions as well as economic and health considerations is effective in reducing disease levels? There are published results based on indicators, such as risk, and reproductive number; or study the effect of risk perception on disease dynamics [e.g. 4–8] that have attempted to answer this question. In [4], geolocalized risk information for COVID-19 is used that gives the probability of finding, at time *t*, at least one infected person in an event of *k* individuals via a binomial assumption of homogeneous risk; [5] revises the 1918 influenza pandemic for which a basic mathematical model is constructed that through a time-dependent transmission contact rate, simulates schools openings and closures, temperature changes, and also changes in human behavior. In particular, changes in human behavior are formulated through risk perception to reported deaths by the disease. In [7] a mathematical model to explore COVID-19 dynamics in Chile is presented. This model includes vaccination dynamics and two perception risk variables, for the unvaccinated and vaccinated population respectively. Here it is assumed that the transmission contact rate depends on the risk in such a way that as risk perception increases, the probability of contagion decreases.

In the present work, we propose a surveillance indicator for a generic acute respiratory infection (ARI) based on epidemiological traffic light policies that integrate the observed disease dynamics, the perception of the risk of contagion, and the implementation of NPIs. We postulate a mathematical model with annual weather-driven cycles, a risk variable, and vaccination dynamics. We use ideas from the classic optimal control theory to formulate a problem that captures the trade-off between epidemiological and economic impact. The resulting policies choose the best action for the next decision period and are represented by a traffic light with four colors, where red represents the more strict policy and green is the less strict one. Our results highlight the role of the interplay between the length of the time-decision period and the size of the population. This work is theoretical in nature but inspired by the patterns and interventions applied to COVID-19 (see Figure 1).

In what follows, we present a mathematical model of a traffic light based on the Kermack-McKendrick equations that accounts for these variables and their temporal dynamics in the presence of an active epidemic of ARIs.

## 2. Mathematical model formulation

We present an extension of the classic Kermack-McKendrick mathematical model with four compartments (see Figure 2). Let the total population *N* be constant. *N* is split into four compartments: susceptible *S*(*t*), infected *I*(*t*), recovered *R*(*t*), and vaccinated *V* (*t*). The disease life-history is the standard for a directly-transmitted disease, in particular ARIs, modeled as follows: susceptible individuals can become infectious through contact with an infectious individual. After a period of time 1/*γ*, infected people recover with waning immunity. Recovered people lose their immunity after a period of time 1/*θ*. Susceptible individuals are vaccinated at a rate equal to *ϕ*. The vaccine is imperfect with efficacy *σ*. Finally, vaccinated individuals that do not acquire the disease, lose their vaccine-induced immunity after a period of time 1/*ω*. Our model also incorporates vital dynamics.

**Figure 2:**
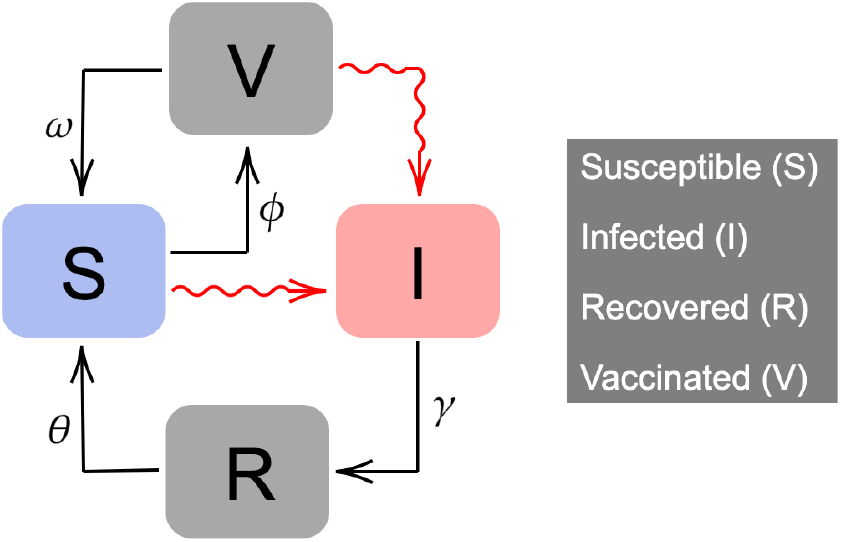
Compartmental diagram of disease transmission dynamics which including vaccination. Red connections represent transmission dynamics which include a risk variable *C*(*t*).

Epidemiological surveillance of the COVID-19 epidemic has generated a large amount of information regarding the dynamics of the indicators used for its control and monitoring, such us the instantaneous reproductive number *R*_*t*_ or the infection risk index developed by [4]. This index gives the probability of finding at time *t* an infected person in a group of *k* individuals. The simple formula is

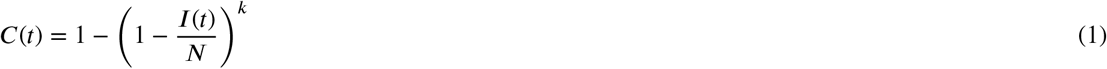

where 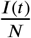 is the number of active cases present in a given locality or region on day *t*. In [4] the number of active cases is computed using the ascertainment ratio derived from data on seroprevalence and contact tracing.

For the force of infection, we postulate equation 2. We incorporate the risk index to introduce human behavior into the transmission process by assuming that the perception of risk conveyed by the knowledge of this index influences transmission dynamics, inducing individual’s behavior averse to getting sick. Clearly, a low-risk index conveys a perception of safety in the population (with the consequent relaxation of mitigation measures, i.e. neglecting socialdistancing) likely triggering an increase in the effective contact rate. A high-risk index produces the opposite effect. With these considerations, the force of infection is given by

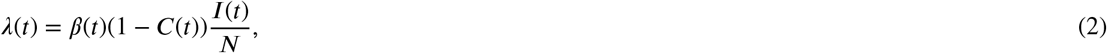

with 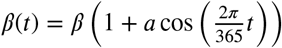 reflecting the seasonal component driving the epidemic.

One important parameter is *k*, the average size of groups where transmission may occur. We see *k* as a control parameter directly related to the perception of risk and risk avoiding behavior of an average individual [4]. Health authorities may recommend, depending upon the observed state of the epidemic, either to reduce the size of the groups (reduce *k*) or relax the mitigation measures and allow large gatherings (large *k*). Using the definition of risk (1) and differentiating with respect to the time *t* we obtain the following equation for the risk of contagion. Note that the structure of this equation includes the per capita change in prevalence *I*^′^/*I* (leftmost expression within parenthesis) multiplied by another expression that depends only on the risk *C*:

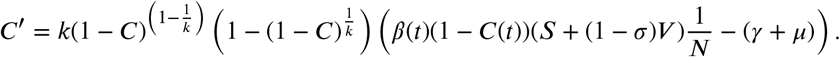

We have now all the components of our mathematical model:

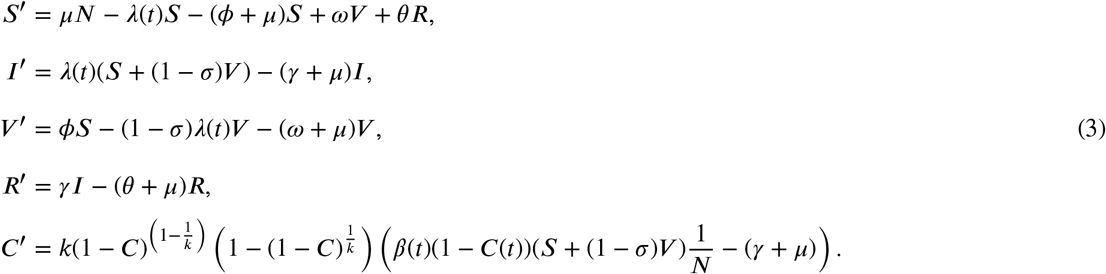

where *λ*(*t*) is given in Equation (2). We normalized system (3) by defining

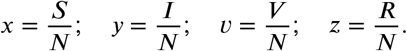

Thus, equations (3) become:

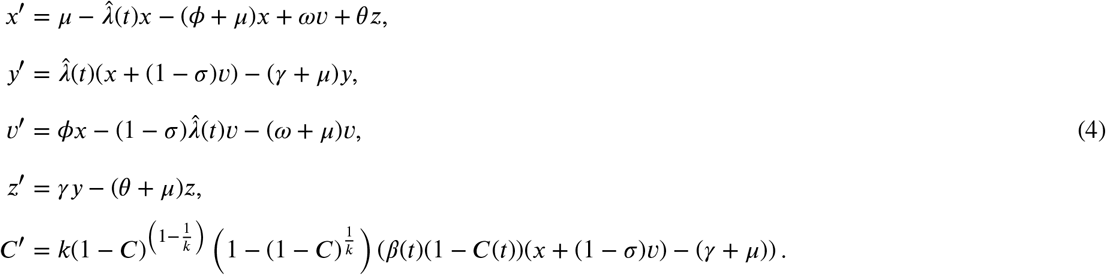

with 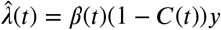. Model (4) is defined in the positively invariant region

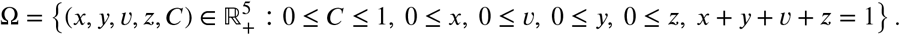

### 2.1. Reproductive Number

Asuming *β*(*t*) = *β*_0_ a constant for small *t*, we calculate the basic reproduction number (*R*_0_), given by

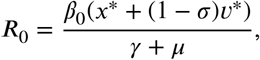

where *x*^*^, *v*^*^ represent the equilibrium values at disease-free equilibrium with (clearly, we have *C*^*^ = 0):

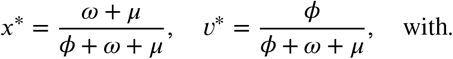

The parameter *β*_0_ is the constant contact rate at the beginning of the epidemic and *β*(*t*) is the time-varying contact rate. Thus, the effective reproduction number at each time is

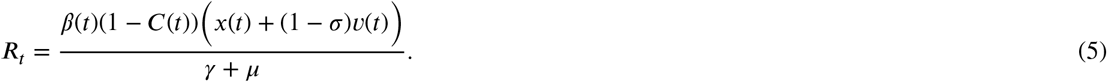

Note that this is a modification of the usual time reproduction number that now includes the risk index. Thus, if *C* is low *R*_*t*_ increases and it decreases if *C* is high, independently of the values of *x* and *v*, the susceptible and vaccinated population proportions, respectively.

## 3. Traffic-Light Policies and Best Response Optimal Control

Individual behavior has played a major role in the evolution of the COVID pandemic [9–12] as it has been with other respiratory diseases. In particular, during the COVID-19 pandemic, human behavior played a significant role in vaccine acceptance and for adoption of NPI [13–16]. Social distancing has been a widespread measure to control and mitigate COVID-19 but also an effective measure against other respiratory infections such as influenza and RSV, according to the CDC and WHO, as their low prevalence in 2020, 2021 and 2022 has shown. Of the many components of social distancing, we address only one which is the limitation of individual’s group size [4]. Our view for a mitigation strategy focuses both on the number of individuals attending an event and the current prevalence as an indicator of risk of contagion. In principle, by controlling the size of meetings, a policy can mitigate transmission and diminish incidence. Obviously, these restrictions have to achieve a balance between the health, the social and the political implications of their implementation. We use the definition of risk of contagion described in [4] because is intuitively clear, simple to explain and straightforward to calculate if one has some estimate of the current ascertainment factor to approximate the actual total number of individuals and thus the true prevalence. Our underlying hypothesis is that the design of a feasible and optimal way to achieve maximum mitigation and control and higher compliance, both transmission and a measure of the risk of contagion have to be considered. In economic terms, the efficacy of NPIs in general, is a trade-off between the cost of its implementation, that might be perceived as interfering with rights, needs and customs, and the health benefit that such implementation conveys.

### Notation

We denote by *u* = *u*(*φ*(*ξ*^⊤^)) the control function on the dynamic-implicit state *ξ*(*t*)^⊤^ = (*s*(*t*), *y*(*t*), *v*(*t*), *z*(*t*), *C*(*t*)). Here *φ* classifies the dynamic state *ξ*^⊤^ in terms of the traffic light colors—green, yellow, orange, red. As we will see below, these colors constitute an objective or at least useful reference for the severity and risk of contagion in a current outbreak. Without loss of generality, and as an example to illustrate our idea, given the number of true active cases at time *t* (the prevalence *y*(*t*)), risk *C*(*t*), and the instantaneous reproductive number *R*_*t*_, we (arbitrarily in this example but ideally based on sound health policy indicatives) classify the initial state of an epidemic outbreak 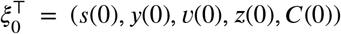 providing several intervals for the main indicators of the traffic-light, which are prevalence, the risk index and the instantaneous reproduction number:

**Green** The prevalence of active cases is less than 10%, the risk index is below 0.3 and *R*_*t*_ remains between [0, 0.7].

**Yellow** The prevalence lies between 10% and 25%, the risk index is above 0.3 but less than 0.5, and the instantaneous reproductive is in the interval [0.7, 1.0).

**Orange** The prevalence is high and above 25% but less than 35%, the risk index lies in the interval [0.5, 0.85) and the *R*_*t*_ is between 1.0 and 2.0.

**Red** The prevalence is greater than 35%, the risk index is above 0.85, and the instantaneous *R*_*t*_ exceeds 2.

For our traffic-light system, the function *φ* maps the initial state *x*_0_ to a specific color

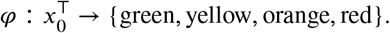

We assume that the decision maker choose a strategy from a finite and well-defined set of actions. This set of actions produces a particular effect associated to a color. The consequence is the modulation of the transmission rate *β* and the size of groups *k*. We define two control functions *u*_*β*_ and *u*_*k*_, and rewrite our model as follows. First, we define the (controlled) infection force and (controlled) group size by

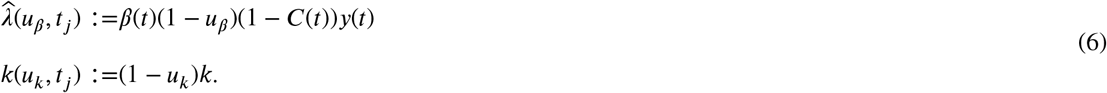

Substituting the above expressions in model (4), we deduce our controlled version.

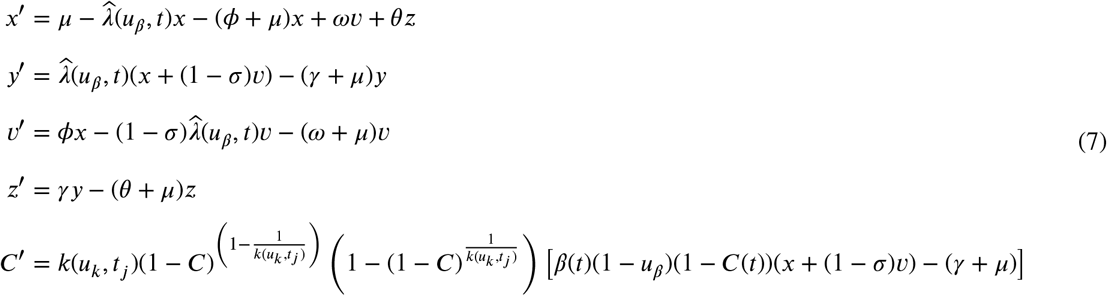

To evaluate the performance of each strategy, we consider the cost functional

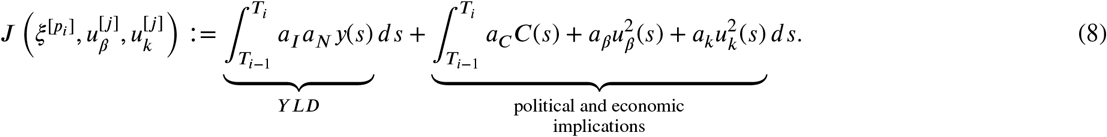

Here *j* denotes one of the action from the admissible set of traffic-light colors {*green, yellow, orange, red*}. The ndex *i* runs over the stages according to the *i*-decision period *p*_*i*_, *i* = {0, …, *M* − 1}. Thus, the implementation of cost and its political and economic impact at each stage follows the linear-quadratic form

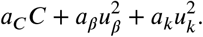

In consequence, we define the mitigation protocol in terms of the color of the traffic-light according to the following criteria:

**Green** Enforce mask wearing and health distance

**Yellow** This set of strategies are preventive and imply mobility restrictions, to reduce the nominal transmission rate *β*_0_ and the meeting-size *k* to 85% of the baseline (no epidemic).

**Orange** This set of strategies aim to more restrictive protocols to quickly reduce prevalence, but allowing enough mobility in an attempt to minimize the impact on economic variables.

**Red** This set of strategies corresponds to an emergency state. Here the infection prevalence is about to exceed the health services capacity. Thus, the decision is aimed to reduce to a minimum the mobility and size of meetings.

#### Hypothesis 3.1

*Let ξ*_0_ = (*x*(0), *y*(0), *v*(0), *z*(0), *C*(0))^⊤^ *the initial state of system* (3). *Thus, according to the above classification, we assign an initial color to the traffic-light controller under the following conditions:*

(H-1) *The decision agent chooses a strategy from the set listed above, thus selecting a color: —{green, yellow, orange, red}*.

(H-2) *The agent makes a decision on the traffic light color every (fixed) number of weeks*.

(H-3) *The decision taken minimizes the cost-functional J* (8) *subject to the dynamics of model* (3).

To perform our simulations, we segment the time horizon interval, [0, *T*] in *M* decision periods such that in each sub-interval [*t*_*i*_, *t*_*i*+1_), *i* = 0, …, *M* − 1, the decision-maker determines the color that minimizes (8). The optimal control problem is designed to produce the best light color change in the decision period *p*_*i*_. In mathematical terms, the optimal control problem for each stage *i* is

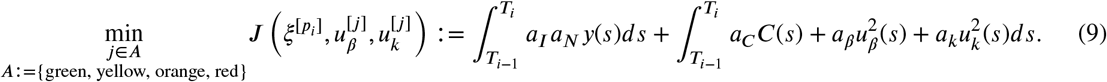

such that

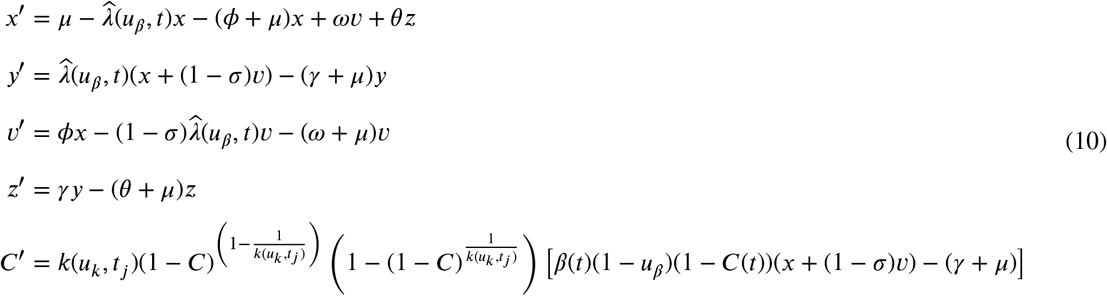

### On the interretation of the functional cost and its weights

Here we describe in more detail the cost functional (8). To quantify the burden of an ARI, we follow the guidelines of WHO [17] for calculating the so-called disability-adjusted life year (DALY). This time-based measure combines years of life lost due to premature mortality (YLLs), years of life lost, and years of healthy life lost due to disability (YLDs). Because our formulation does not consider disease mortality, we only employ the related term to quantify YLD. In mathematical terms, we use *a*_*I*_ *a*_*N*_ *y*(·) to describe YLD concerning a population of size *a*_*N*_ .

We also penalize the changes on risk perception under the hypothesis that it is proportional to the state given by *C*(·) assume that risk perception implies, for example, political costs. For example, if the perception of risk remains high and lasts more than one month, then it would damage the political image of the Health Council and/or government staff. Further, we suppose that applying a restriction o mobility or the size of meetings implies a political-economic cost and this cost follows the quadratic form of the second integral in (8). These terms and our formulation of YLD allow us to quantify and calibrate the trade-off between the health benefit and economic implications. For example, if the decision-maker aims to diminish the cost of implementation, then she would decrease the weights *a*_*β*_, and *a*_*k*_ while increase the cost due to prevalence *a*_*I*_ —or a similar alternative strategy to obtain the same bias—to accordingly balance the trade-off between the health benefit that is quantified by the YLD and the politic-economic implications.

#### Algorithm 1

Best traffic-light policy

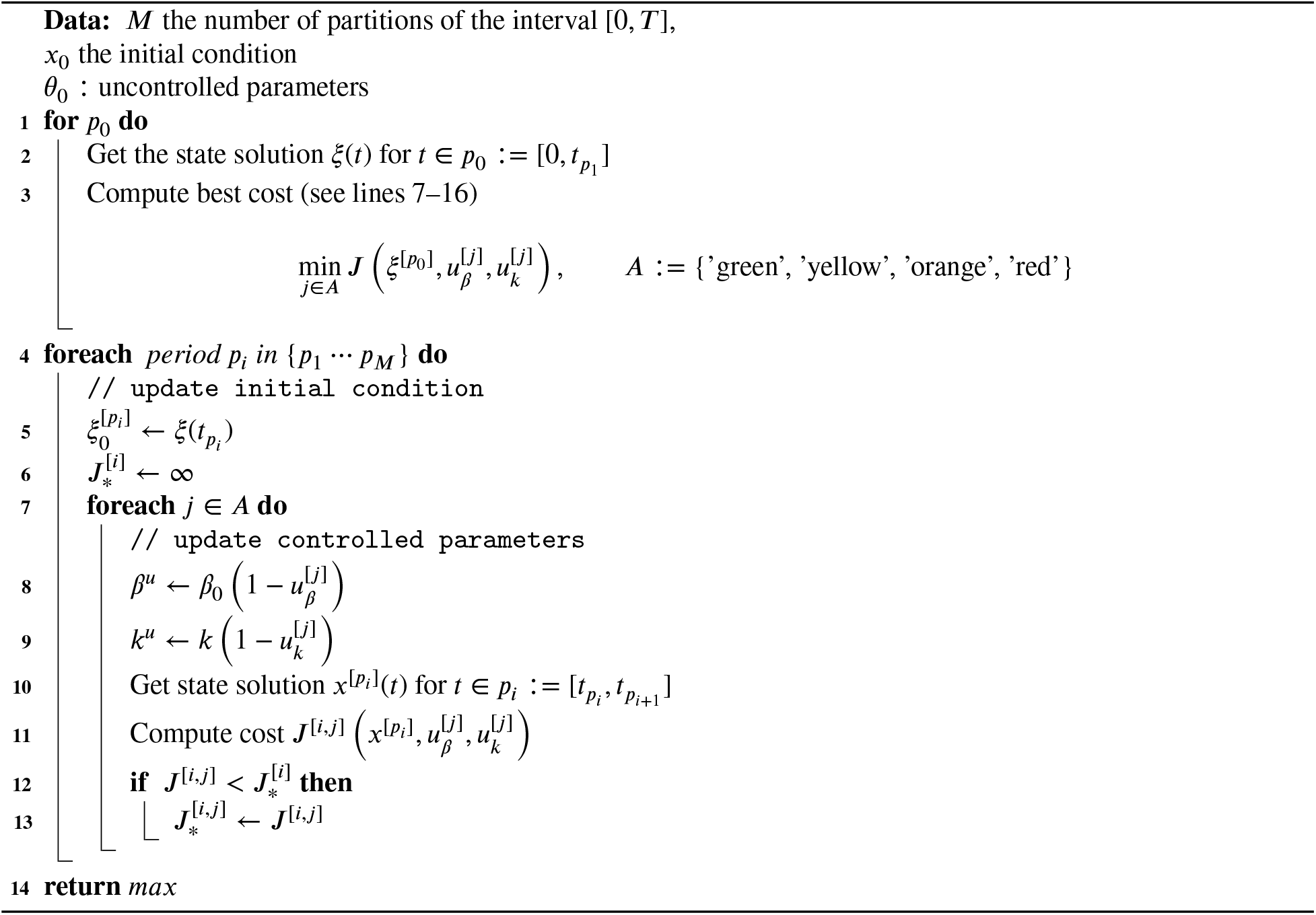

*Implication of different configurations of weights*

## 4. Results

### 4.1. Implementation

In this section, we evaluate the efficiency of the protocol according to the best response defined as the action that minimizes the cost *J* . To this end, we consider a respiratory disease in its starting phase where the number of cases is low but grows as the effective reproductive number increases above 1. Thus, after one period (one week), the decision maker chooses an action consistent with the best traffic-light color—the color that minimizes the cost *J* for the next period. This decision impacts the transmission contact rate *β* and the size of meetings *k*. In the following period, the decision maker again evaluates the epidemic state and takes a new action. This process continues until the simulation reaches *M* periods. To begin these simulations, we assume that there are two infected individuals at the beginning of this study. Table 1 shows the parameter values used in the simulations that follow.

**Table 1.** Baseline parameter’s value of system (10). ^*^ Fixed value for all experiments. ^†^ Population size *N* change according to the experiment in Figure 8. ^‡^ Fixed value for the remaining figures.

| Symbol | Value | Unit |
| --- | --- | --- |
| **Baseline\*** |  |  |
| $\mu$ | $3.913\,894 \times 10^{-5}$ | day<sup>-1</sup> |
| $\gamma$ | 1/10 | day<sup>-1</sup> |
| $\phi$ | 0.002 | day<sup>-1</sup> |
| $k$ | 200 | inhabitants |
| $\omega$ | 1/180 | day<sup>-1</sup> |
| $a$ | 1 | dimensionless |
| $\theta$ | 1/180 | day<sup>-1</sup> |
| $\beta$ | 0.3 | dimensionless |
| $\sigma$ | 0.95 | dimensionless |
| **Initial conditions†** |  |  |
| $\xi_0 = (x_0, y_0, v_0, z_0, C_0)^\top$ | $\left( \frac{N-2}{N}, \frac{2}{N}, 0, 0, 1 - \left(1 - \frac{2}{N}\right)^k \right)$ | population – distribution |
| $a_I$ | 0.0001 | dimensionless |
| $a_C$ | 0.001 | dimensionless |
| $a_k$ | 0.001 | dimensionless |
| **Figure 6** |  |  |
| $a_\beta$ | 0.005‡, 0.03, 0.001 | dimensionless |
| **Figure 7** |  |  |
| $p_i$ | 7 ‡, 14, 28 | days |
| **Figure 8** |  |  |
| $a_N, N$ | 1×10<sup>6</sup>, 1×10<sup>5</sup> ‡, 5×10<sup>4</sup> | inhabitants |

Figure 3 compares the dynamics of an uncontrolled outbreak and the dynamics of the disease controlled by the traffic light. In our example, the best traffic-light policy response suggests staying in green when prevalence is low, and risk remains under 20%. In this scenario, the transition from green to red occurs when prevalence and risk are sufficiently large and with a relatively high *R*_*t*_. The Figure shows how this action “flattens” the curve, and allows the best response to change to orange. Later, the reduction in prevalence forces a transition to yellow and finally to green. Then for a best response we obtain the sequence from green to red, red to orange, orange to yellow, and yellow to green. As a consequence, prevalence, and risk decrease, even though *R*_*t*_ has kept a similar magnitude both in the observed and in the uncontrolled dynamics.

**Figure 3:**
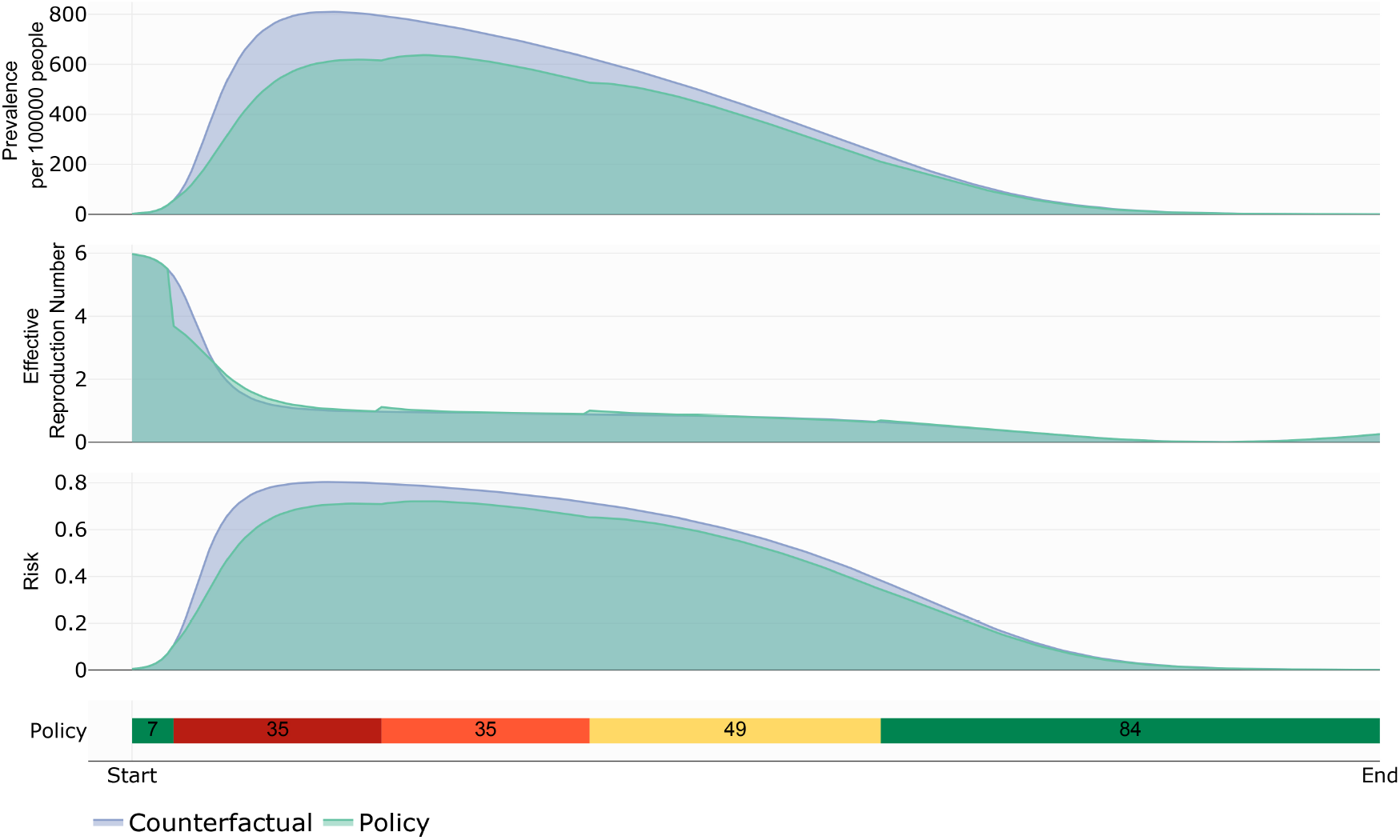
Prevalence, instantaneous reproduction number, risk index and light traffic policy according to the best response. Cost functional parameters are equal to *a*_*I*_ = 0.0001, *a*_*N*_ = 100 000, *a*_*β*_ = 0.05, *a*_*k*_ = *a*_*c*_ = 0.001.

Figure 4 extends the time horizon illustrated in Figure 3. This case suggests that to obtain the best response, certain common features exists requiring a green light at the beginning and end of the outbreak, and orange and yellow lights near the outbreak peak.

**Figure 4:**
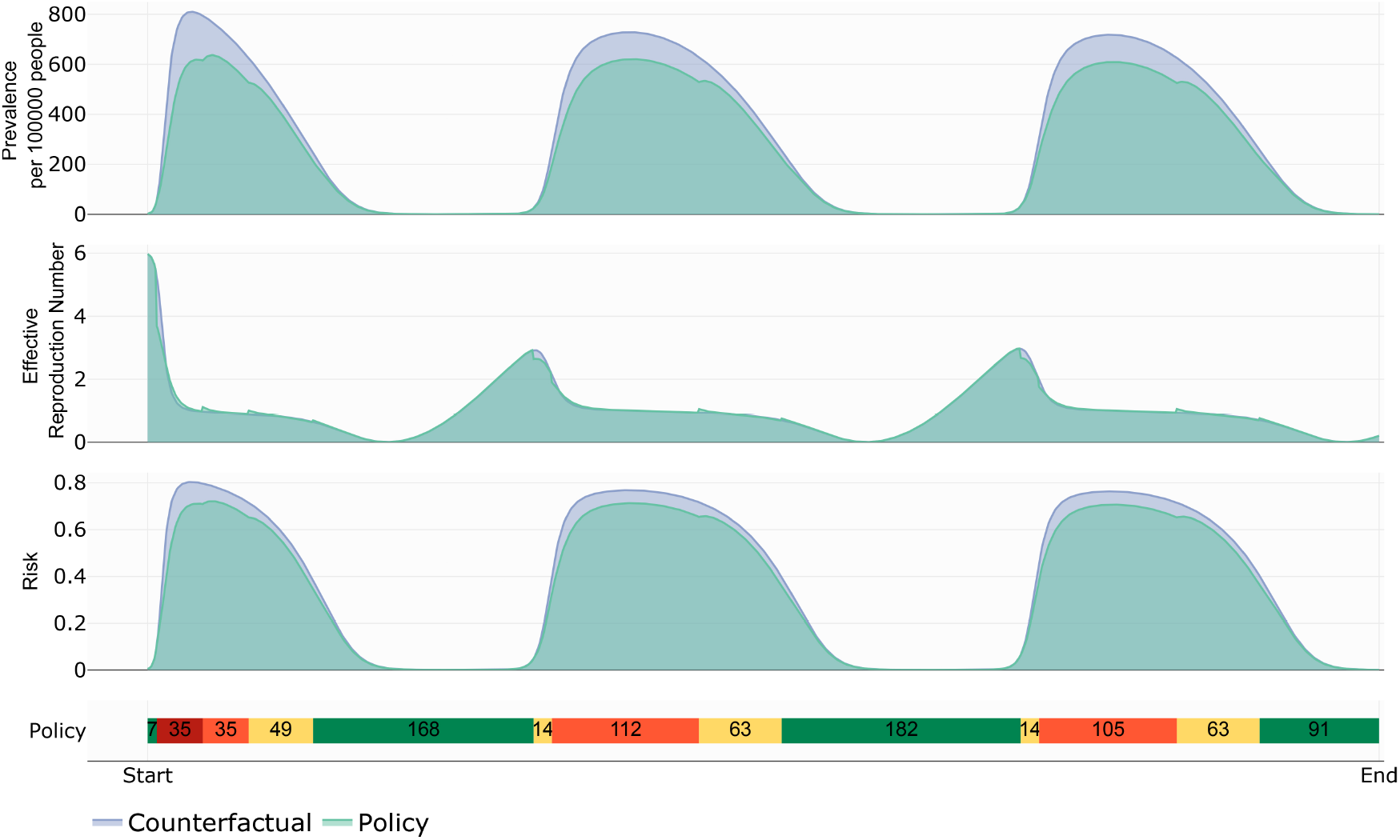
Counterfactual vs. controlled dynamics: Prevalence, *R*_*t*_ and risk. Cost parameter values are equal to *a*_*I*_ = 0.0001, *a*_*N*_ = 100 000, *a*_*β*_ = 0.05, *a*_*k*_ = *a*_*c*_ = 0.001.

Figure 5 illustrates the dynamics of the cumulative incidence and the cost functional. By the end of the simulation time, the optimal policy achieves a reduction in cumulative incidence and the associated cost of more than 10% and 15%, respectively, in contrast to the counterfactual scenario.

**Figure 5:**
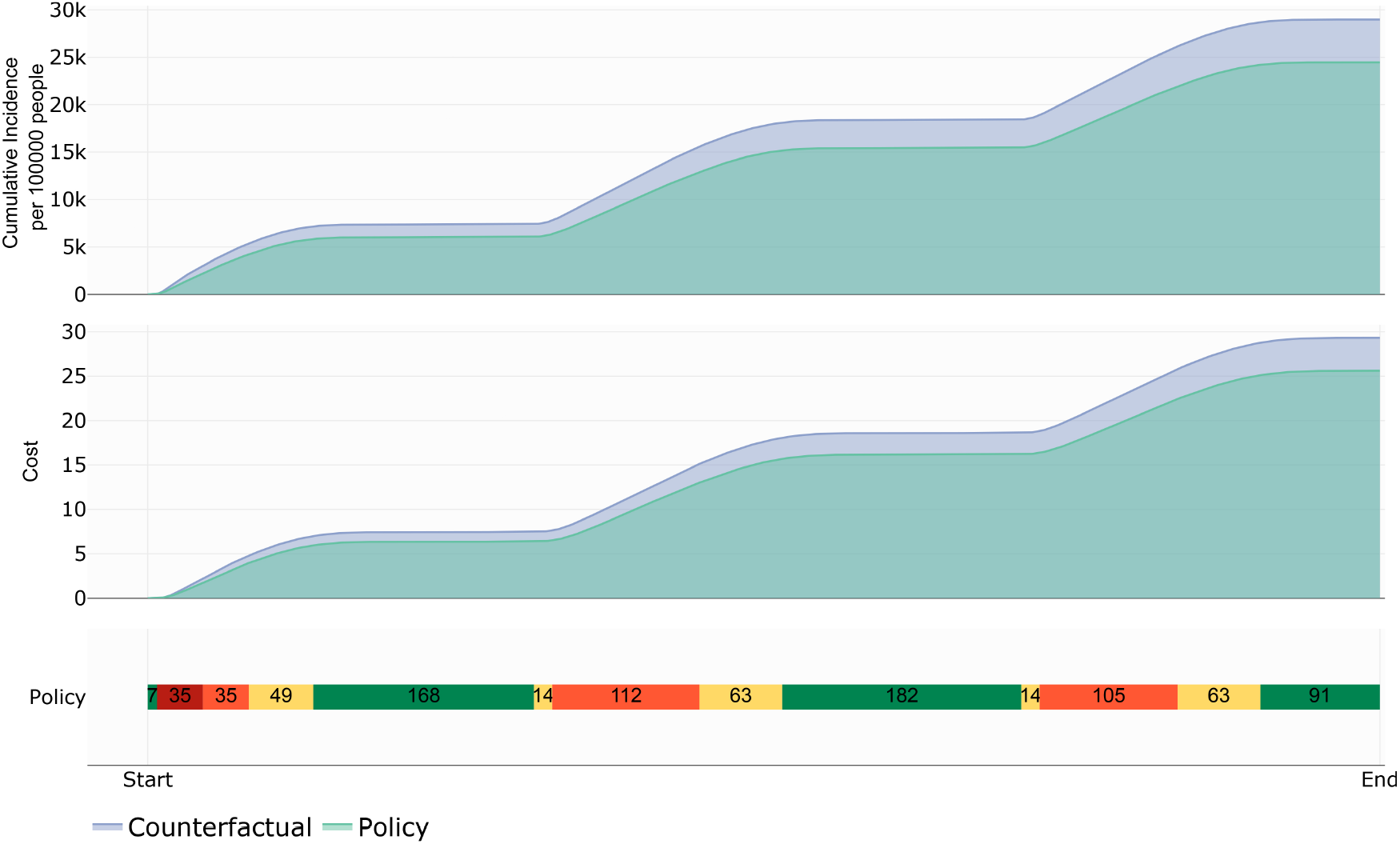
Counterfactual vs. controlled dynamics: Cumulative incidence and Cost. Cost functional parameters are *a*_*I*_ = 0.0001, *a*_*N*_ = 100 000, *a*_*β*_ = 0.05, *a*_*k*_ = *a*_*c*_ = 0.001.

### 4.2. The role of cost, decision period and population size in the traffic light design

Figure 6 illustrates how changes in the cost parameters produce changes in the traffic light. We compare non controlled dynamics with several policies with different *a*_*β*_ for a decision period of one week. Table 1 displays the other parameter values being used. Observe that if *a*_*β*_ increases, then the corresponding policy is less strict. Figure 6 shows that our projected policies indeed reduce prevalence levels. Furthermore, as the policy is more strict, the prevalence reduction will be higher. This last observation is apparent in policy 3 for which the traffic light is mostly red.

**Figure 6:**
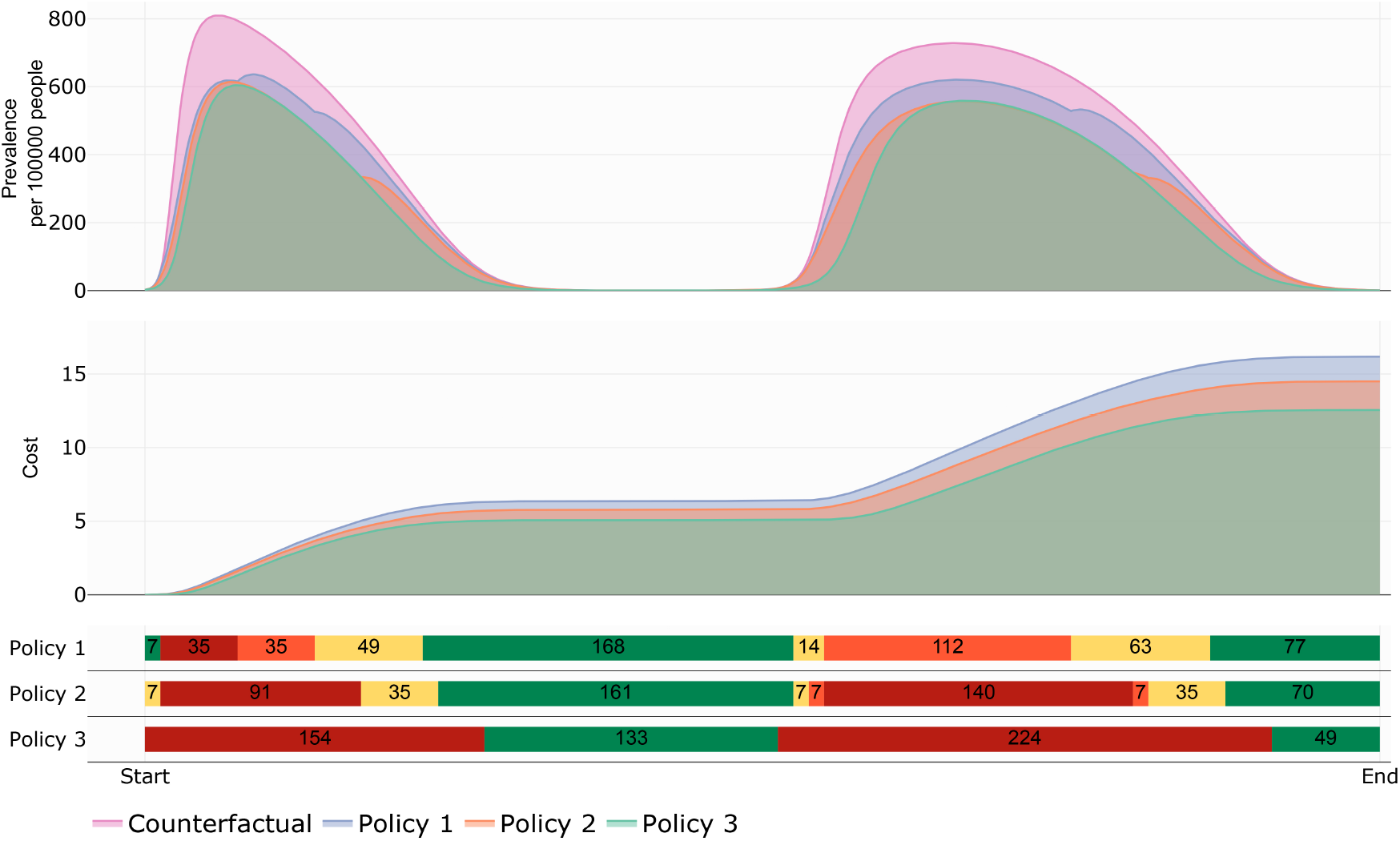
Transmission dynamics when varying *a*_*β*_ . Policy 1: *a*_*β*_ = 0.05. Policy 2: *a*_*β*_ = 0.03. Policy 3: *a*_*β*_ = 0.001. Other cost parameters values are *a*_*N*_ = 100 000, *a*_*I*_ = 0.0001 and *a*_*k*_ = *a*_*c*_ = 0.001. The decision period is equal to one week.

Figure 7 exemplifies the importance of a well-chosen decision period. We contrast non-controlled dynamics against different policies. These policies are obtained using decision periods equal to one week (Policy 1), two weeks (Policy 2), and four weeks (Policy 3). Other parameters are given in Table 1. We observe that depending on the length of the decision period the policy can be more or less strict. For example, if decisions are taken every four weeks, the epidemiological traffic light starts in red and continues without change for 112 days. In contrast, if decisions are taken each week the resulting policy is more flexible. In this case, the epidemiological traffic light starts in green and alternates between green, red, orange, and yellow for the first 112 days. In any case, all policies reduce the prevalence level. Finally, although policy 1 is the less restrictive, we interpret it as being also the less beneficial or effective than the others because the prevalence reduction it achieves is the lowest of all alternatives; it also the most expensive in term of economic costs. In other words policy 1 favors the health benefit over the economic implications.

**Figure 7:**
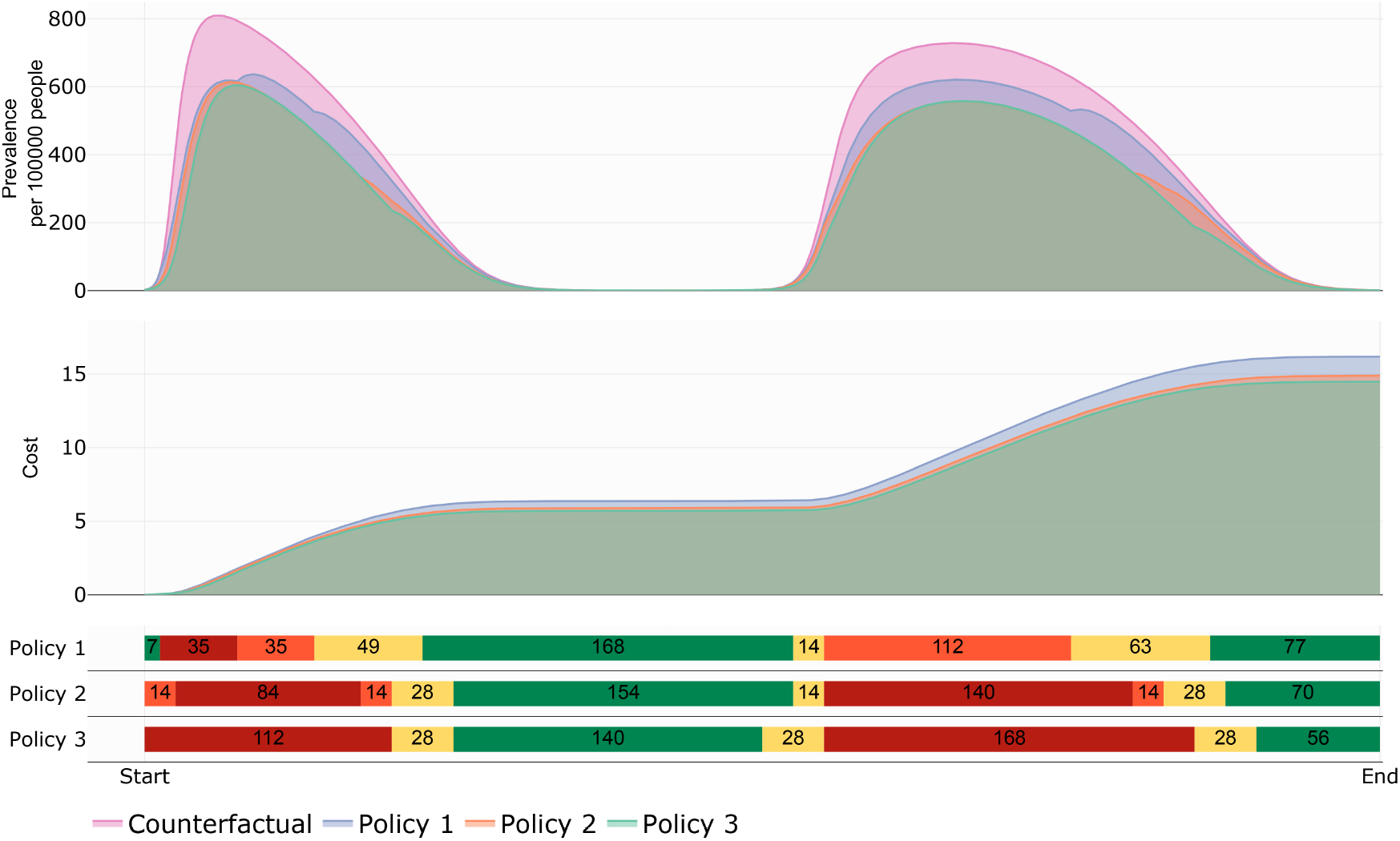
Importance of a well-posed decision period. Each policy is obtained when considering one week (Policy 1), two weeks (Policy 2), and four weeks (Policy 3). Cost parameter values are equal to *a*_*I*_ = 0.0001, *a*_*N*_ = 100 000, *a*_*β*_ = 0.05, *a*_*k*_ = *a*_*c*_ = 0.001.

Figure 8 shows the role of population size *N* in the traffic light design on the total cost. In Figure 8 policies 1, 2, and 3 correspond to population sizes equal to 1 000 000, 100 000, and 50 000 inhabitants, respectively. Parameter values are equal for all policies. Observe that policy 1 renders a traffic light protocol with predominantly red color, while policy 3 renders color green as the dominant one. Note too that the total cost of applying the policy to the largest population (policy 1) is more than 10 times higher than the policy with the smallest population (50 000 inhabitants). However, if we consider the per capita cost, then policy 3 (50 000 inhabitants) is the one with higher cost.

**Figure 8:**
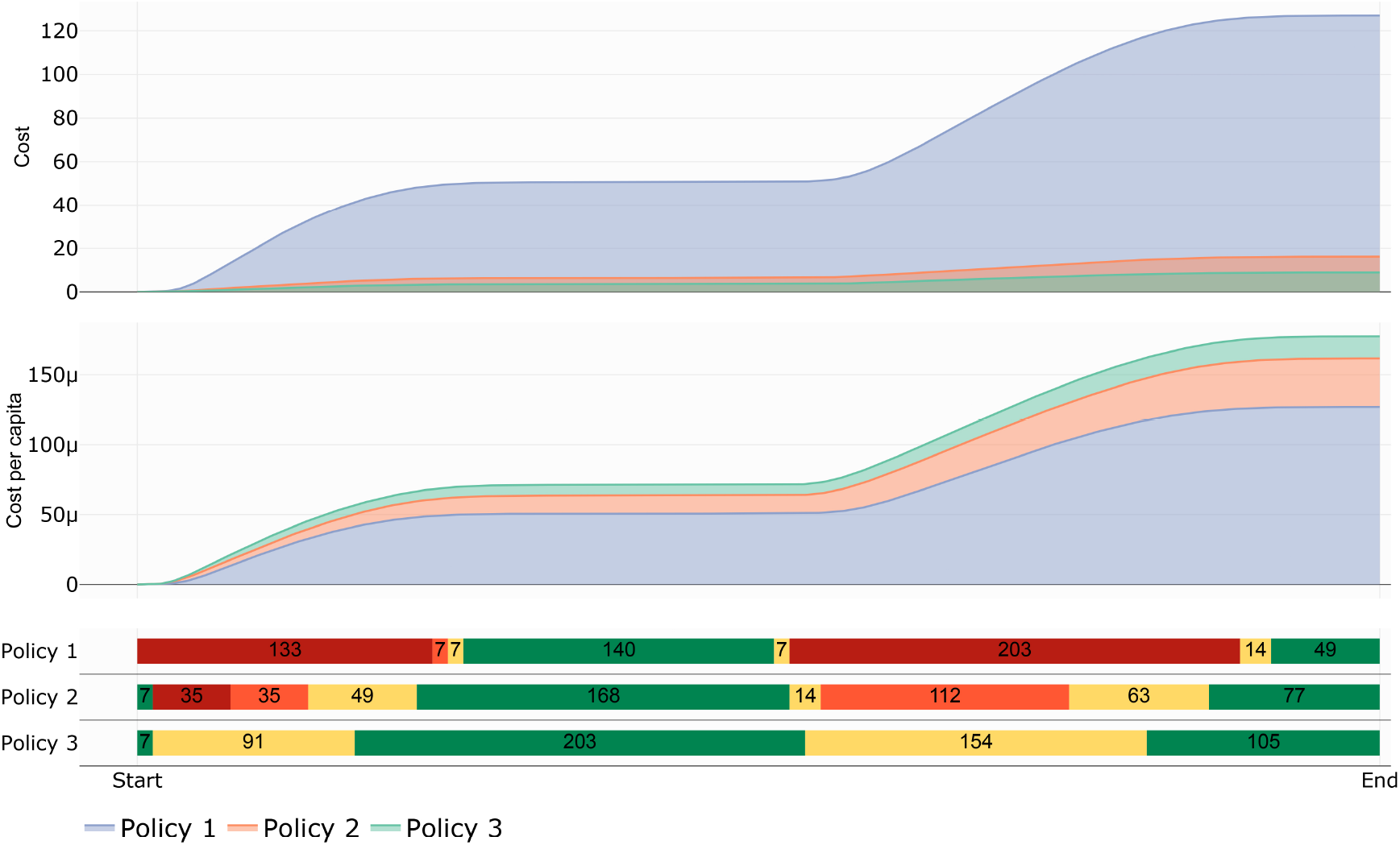
Impact of the population size on the cost and the light traffic policy. Each policy is obtained when considering *a*_*I*_ = 0.0001, *a*_*β*_ = 0.05, *a*_*k*_ = *a*_*c*_ = 0.001, and the decision period is equal to one week. Policy 1 represents the dynamics for 1 000 000 inhabitants (*a*_*N*_ = 1 000 000). Policy 2 illustrates the dynamics for 100 000 inhabitants (*a*_*N*_ = 100 000). Policy 3 represents the dynamics for 50 000 inhabitants (*a*_*N*_ = 50 000).

## 5. The impact of vaccination on the traffic light design

In this section we look at our traffic light designs assuming that vaccines are available and the population is being immunized. Our models mimics a general acute respiratory infection and, certainly, there are no vaccines for all diseases or, if they exist, coverage might not be very large or availability may be restricted. COVID-19 was a clear example of the above. Vaccines were administrated about a year after the pandemic started and when the vaccine became available there were not enough to apply to the entire population. This is the reason why we consider it important to explore the role of vaccination dynamics in our traffic light design.

Figure 9 illustrates the impact of vaccine availability (either limited vaccine application or lack of vaccine) on costs and traffic-light colors. Simulations were run for a time equal to 80 weeks. Policy 1 represents a scenario without vaccination; olicy 2 illustrates a scenario without vaccination in the first year; policy 3 shows a scenario with vaccines available immediately. We observe that the faster the vaccination is implemented, the less strict policies need be. For example, policy 3 stays only five weeks in light-color red, in contrast to policies 1 and 2 that stay 23 and 21 weeks in that color, respectively.

**Figure 9:**
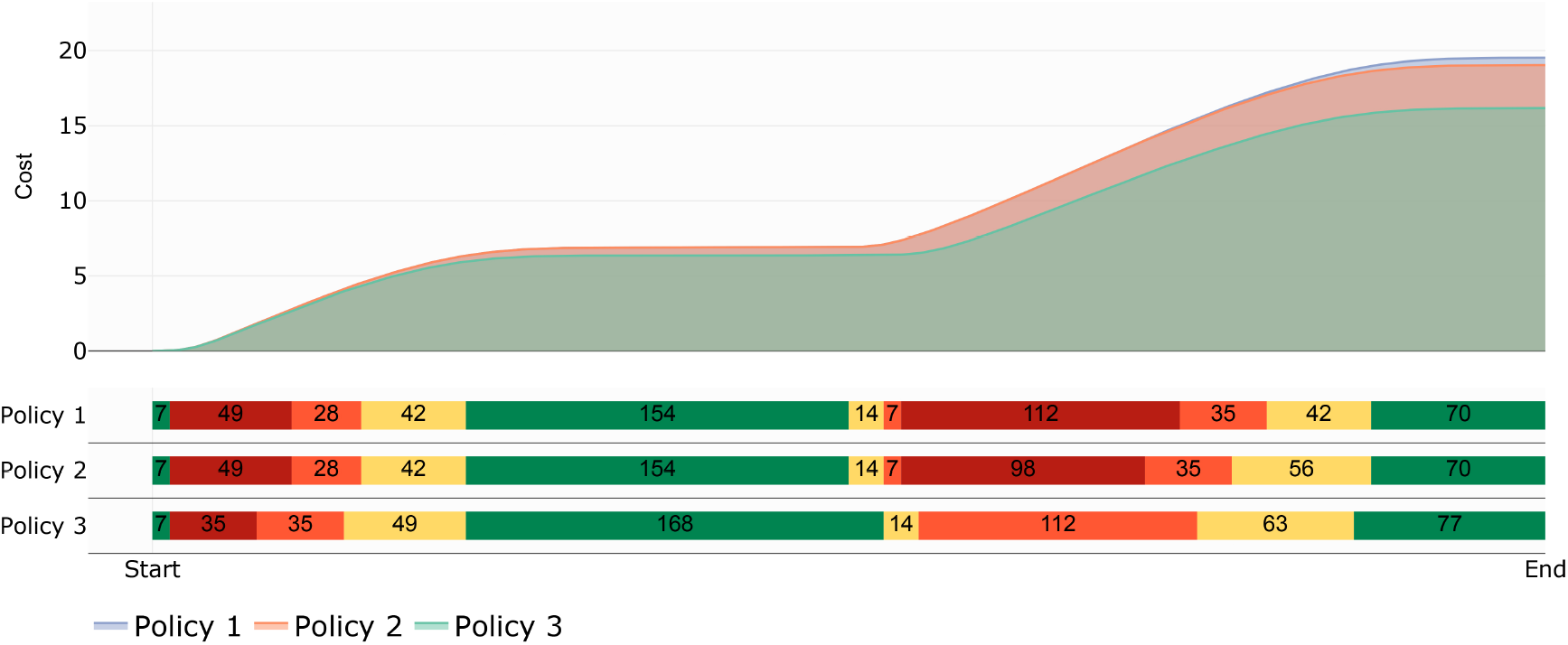
Impact of vaccine availability on the cost and light traffic policy. Each policy is with *a*_*I*_ = 0.0001, *a*_*N*_ = 100 000, *a*_*β*_ = 0.05, *a*_*k*_ = *a*_*c*_ = 0.001, and the decision period is equal to one week. Policies 1, 2, and 3 are obtained for the scenarios without vaccination, without vaccination available after the first year, and with vaccination all the time, respectively.

Figure 10 shows the impact of the vaccination strategy on the cost and the policy. Observe that the larger the target population to vaccinate is, the less restrictive the policies will be. Policies 1, 2, and 3 represent scenarios in which the target population to vaccinate in 28 weeks, is close to 2%, 32%, and 54%, respectively. We observe that the scenario with lowest target population renders a more strict policy. On the other hand, even though policies 2 and 3 are similar, the total cost exhibits a reduction close to 16% in the former compared to the latter.

**Figure 10:**
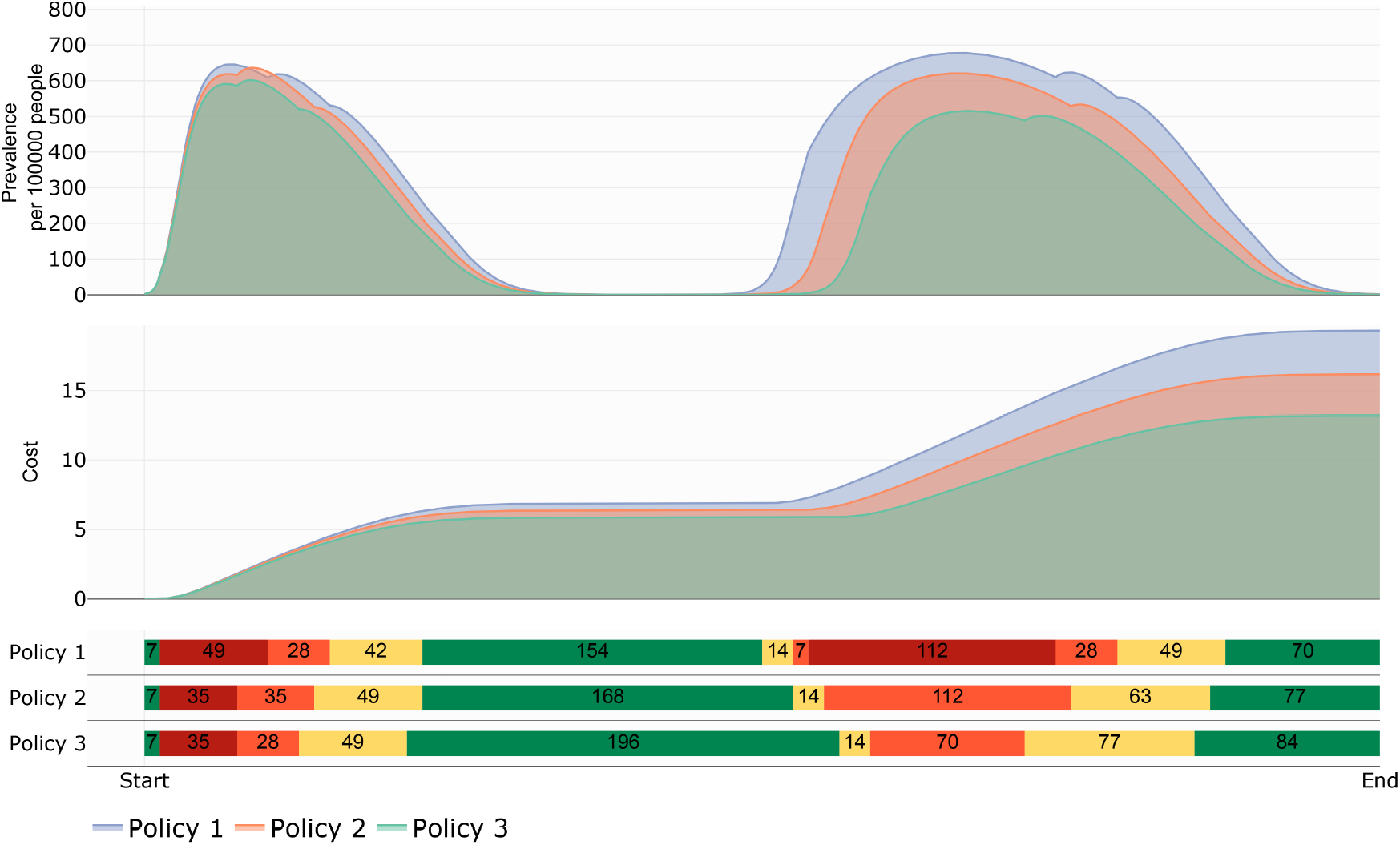
Impact of the vaccination strategy on the cost and the light traffic policy. Each policy is obtained when considering *a*_*I*_ = 0.0001, *a*_*N*_ = 100 000, *a*_*β*_ = 0.05, *a*_*k*_ = *a*_*c*_ = 0.001, and the decision period is equal to one week. Policy 1, 2, and 3 are obtained when vaccination rate (*ϕ*) is equal to 0.0001, 0.002, and 0.004, respectively.

## 6. Discussion

Given a region, city or town with a given population size, we have developed a theoretical model to evaluate the impact of risk perception and the associated behavior and epidemic dynamics. Behavioral effects are notoriously difficult to model, and we have attempted to approximate this phenomenon using a dynamic model that incorporates a measure of risk perception. In particular, we focus our work on the perception of risk associated to the size of events that people attend, adapting a risk index that measures the risk of contagion when attending meetings of certain size. With this model, we design and evaluate the optimal best response to NPIs mitigation measures that may include protocols for mobility restriction, mask wearing, social distance and others. Our model describes then the traffic-light associated to risk perception and the underlying epidemic dynamics and measures their impact on policy decisions implemented by a hypothetical decision maker. Although our model is simple, it is able to capture the impact of increasing levels of NPIs interventions in the presence of vaccination and also it can provide a measure of the economic cost associated by a given strategy. Our simulations suggest that the frequency at which implementation decisions are made and the cost weights used, modulate the strictness of the light-traffic changes. We found that a sequence of red and green colors is the best to control prevalence when the decision maker values the health benefits of the population over the direct economic impact. We also observe that this strictness is closely related to the propers of the cost functional—the mean cost contribution per inhabitant and the weights *a*_*I*_, *a*_*β*_, *a*_*k*_, *a*_*C*_ . These weights respectively represent the importance given to prevalence, NPI’s, group size and risk perception. In other words, the decision maker can adjust these weights according to the cost given to the size of meetings, or the political, economic and cultural costs measured by *a*_*k*_, and *a*_*β*_ . This form of quantify the cost allows us to discern between more convenient policies and driven by a given aim. Therefore, we conclude that the trade-off between health benefits and the cost of economy implications, it is advisable, whenever possible, to identify the best response based of these both aspects. Risk perception has been explored in the literature. Gutierrez-Jara et al. [7] and Lin et al. [6] have published models for covid incorporating different metrics of risk description in their formulation, but with different algebraic structure and aims, compared to ours. In [7], a model is developed to assess the interaction between vaccination and perception of risk concluding that the highest waves of COVID-19 contagion in Chile were related to the risk perception of people. Lin et al. [6] model the course of the COVID-19 outbreak in Wuhan and describe the public perception of risk associated to prevalence incorporating deaths, but do not control its dynamics.These authors propose a similar force of infection to ours, considering too a risk perception variable; however, the coupling of risk with the disease dynamics is different assuming that risk increases or decays as the infectious population increases or decreases. In contrast, we use the risk index proposed by Chande et al. [4] and turn it into a dynamic variable with respect to time. This risk index employs data from the infected population and the size of groups where people meet.

We have here developed a theoretical framework to evaluates the risk and design mitigation policies based in the political perception and health benefits.

Our results show that a careful enforcement and relaxation of restrictions amplify their associated public health benefits and diminish the political and/or economic costs and implications. We show that social variables like risk perception and mobility can be used to advantage in the implementation of mitigation policies for ARIs.

We are aware our model is relatively simple for the transmission dynamics of a complex disease such as an acute respiratory infection where we can have several classes of infectious individuals, presymptomatic stages and so forth, and that there is more than one way to formulate a mathematical model to describe its dynamics. However, our aim has been to evaluate the plausible efficacy of a traffic light system in a simpler framework.

## Data Availability

All data produced in the present study are available upon reasonable request to the authors

## Acknowledgments

JXVH acknowledges support from grants UNAM PAPIIT IN115720 and IV100220. JXVH developed part of this work during his sabbatical leave at the Simons Institute, University of California, Berkeley with partial support from a PASPA-UNAM fellowship.

## Code availability

The code for reproducing all figures is available at the GitHub repository https://github.com/SaulDiazInfante/COVID-19-ControlByaRiskIndex.git.

## Notes

### Competing Interest Statement

The authors have declared no competing interest.

### Funding Statement

This study did not receive any funding

